# Assessing skin temperature over time in a cohort of lower limb cellulitis: A methods comparison study

**DOI:** 10.1101/2024.12.18.24319232

**Authors:** Elizabeth LA Cross, Martin J Llewelyn, A Sarah Walker, Gail N Hayward

## Abstract

**Importance:** Skin temperature assessment is essential for the initial diagnosis of cellulitis and monitoring treatment response. Currently, this is subjective and can contribute to overdiagnosis.

**Objective:** To characterise skin temperature changes over time in cellulitis and compare two more objective measurement approaches, a thermal imaging camera (TIC) and a non-contact infrared thermometer (NCIT).

**Design:** A methods comparison study nested within a prospective cohort. We measured limb temperatures daily for four days using a TIC and two NCITs (2-4 measurements/time points).

**Setting:** Two acute hospitals in the United Kingdom’s National Health Service.

**Participants:** Adults (age ≥18 years) diagnosed with lower limb cellulitis who attended hospital for antibiotic treatment.

**Main Outcome(s) and Measure(s):** We used linear mixed-effects models to quantify changes in temperature over time and intraclass correlation coefficients (ICC) to assess reliability. We compared temperature measurements between devices using Lin’s concordance coefficients and Bland-Altman plots with estimated 95% limits of agreement (LOA).

**Results:** 202 patients were included: 95% white ethnicity. The affected limb remained hotter than the unaffected limb across all days. Baseline affected limb temperatures varied between 33.1-36.9°C and limb temperature differences between 2.4-3.4°C, depending on the device. All devices showed significant reductions in affected limb temperature per day, with the largest decrease for the TIC (−0.34°C per day, 95%CI −0.48 to −0.19, P<0.001). Only the TIC and NCIT-1 showed significant reductions in limb temperature difference per day.

All devices had excellent reliability (ICCs≥0.98). The TIC recorded, on average, affected limb temperatures that were lower than NCIT-1 and NCIT-2, by −2.52°C (95% LOA −5.47 to 0.43) and −4.67°C (95% LOA −6.53 to −2.82), respectively. The largest mean differences and the lowest Lin’s concordance coefficient were observed between the TIC and NCIT-2. The NCIT-2 also demonstrated evidence of proportional bias.

**Conclusions and Relevance:** NCIT-2’s poorer performance suggests different NCITs cannot be used interchangeably. Neither the TIC nor NCIT-1 were clearly superior. More advanced analyses of thermal images could prove helpful. Future research should confirm our findings in different skin tones and aim to determine the clinical utility of potential earlier diagnosis or indications of therapeutic failure that thermal imaging might offer.

**Key Points:** *Question:* Should thermal imaging cameras (TICs) or non-contact infrared thermometers (NCITs) be used to measure skin temperature over time in lower limb cellulitis?

*Findings:* In this cohort of 202 adults, absolute affected limb temperature measurements varied widely between devices, including the two NCITs, but limb temperature differences were more similar. The TIC recorded the largest reduction in affected limb temperature over time (−0.34°C per day).

*Meaning:* NCITs’ measurement capabilities differ widely, so these devices cannot be used interchangeably. Due to this and the potential benefits of advanced thermal image analysis, TICs should be prioritised for further study in cellulitis.

## Introduction

Cellulitis is a common bacterial skin infection characterised by warmth, pain, swelling, and acute colour change of the affected skin.^1^ Skin temperature assessment is essential for both the initial diagnosis of cellulitis (to differentiate from mimics such as varicose eczema and lipodermatosclerosis) and for monitoring response to antibiotic treatment.^2,3^ In current practice, this is a subjective clinical assessment and is likely to be very unreliable, especially when conducted by different clinicians over time. One study showed that even substantial temperature differences in extremities of >3°C were only detected by clinicians 76% of the time.^4^ Unsurprisingly, therefore, cellulitis is both overdiagnosed^5,6^ and overtreated, with 30-50% of patients experiencing unnecessarily prolonged antibiotic treatment.^7–9^

Technological solutions that provide objective assessment of skin temperature can potentially improve diagnostic accuracy in cellulitis, thus improving patient outcomes, reducing unnecessary antibiotic treatment and associated harms, including antibiotic resistance, and reducing healthcare costs. Two broad approaches have been applied: non-contact infrared thermometers (NCITs)^10–12^ and thermal imaging cameras (TICs).^13–18^ Diagnostic studies using these devices in cellulitis have found significant temperature differences between affected and unaffected limbs.^13–16,18^ However, few studies have monitored temperatures beyond the point of diagnosis,^10,12,17^ and none have attempted to compare these two technologies. Therefore, the objective of our study was to characterise skin temperature changes over time in cellulitis and compare these two approaches.

## Methods

### Ethics

This study was approved by the East of Scotland Research Ethics Service, Research Ethics Committee (21/ES/0048). All participants provided written informed consent.

### Study design and population

This methods comparison study of two technological approaches (one TIC and two NCITs) was nested within a prospective cohort study of patients with cellulitis conducted between June 2021 and March 2023.^19^ The study was conducted at two acute hospitals in the United Kingdom’s National Health Service (NHS): a large tertiary referral hospital and a district general hospital, both within University Hospitals Sussex NHS Foundation Trust.

Adults (age ≥18 years) diagnosed with lower limb cellulitis requiring antibiotic treatment were eligible for inclusion. The main exclusion criteria were having received three or more calendar days of antibiotics from the hospital for cellulitis or having been treated for a previous episode in the preceding 28 days. Further exclusion criteria are detailed in **supplementary materials p2**.

### Devices

Two devices were evaluated throughout the whole study.

TIC) FLIR ONE® Gen 3 - Android USB-C (Teledyne FLIR, USA), a TIC that attaches to a smartphone with an object temperature range of −20 to +120°C and a reported accuracy of ±3°C.

NCIT-1) Extech® IR200 (Extech Instruments Corporation, USA), an NCIT with a surface temperature range of 0 to 60°C and reported accuracy ±0.8°C.

A third device became available in the study at month 9 and was used on 103 (51%) study patients.

NCIT-2) Thermofocus® 0800/H5 (Tecnimed s.r.l., Italy), an NCIT with a measuring range of 1.0 to 55.0°C and a reported accuracy of ±0.2°C to ±1.0°C, dependent on the measuring temperature and least accurate at extremes of range.

### Procedures

Temperature measurements were taken at the point of maximal temperature on the affected limb and at the corresponding point on the non-affected limb to allow calculation of temperature difference (affected minus unaffected limb temperature) (details in **supplementary materials p2**).

To calculate reliability, repeated measurements were taken from both the affected and unaffected limbs (two measurements for the TIC and three for the NCITs, because a priori it was hypothesised that measurements from the NCITs would be more variable, and taking another repeat measurement added negligible extra time for these devices (<10 seconds) in contrast to the ~2 minutes for each TIC reading and image upload). Temperature readings were made approximately 10 minutes after removing any clothes or dressings. The devices were held at room temperature for at least 10 minutes before readings were taken. Measurements were taken indoors in temperature-regulated clinical areas. Temperature measurements were not provided to treating clinicians.

Where possible, temperature measurements were performed on all patients daily for four days beginning on day 0, defined for the study as the date the patient began their hospital-associated antibiotic treatment for cellulitis (61 (30%) were already taking antibiotics prescribed in the community for a median 3 days (IQR 2,4), in which case day 0 was when the prescription was changed in hospital). Where patients were enrolled after day 0, temperature readings were only available from enrolment. Where patients were discharged before day 3, readings were only available until discharge.

### Statistical Analysis

#### Skin temperature over time

For each device, linear mixed-effects models were used to quantify the mean day 0 temperature and daily change in affected, unaffected, and limb temperature difference, with correlated participant-level random effects for baseline and daily change. Conditional on these random effects, repeated measurements taken within each participant on each specific day were considered independent.

#### Device comparison

To assess reliability, intraclass correlation coefficients (a reliability index that measures the degree of correlation and agreement between measurements) were calculated using a one-way random-effects model to assess the absolute agreement of repeated measurements. To assess repeatability (defined as the consistency of measurements when taken repeatedly by the same device under the same conditions), the repeatability coefficient was calculated using the “REPEATABILITY” Stata module,^20^ estimating 95% confidence intervals from 1000 bootstrap samples. Due to the late introduction of NCIT-2 into the study, we performed a sensitivity analysis comparing the repeatability coefficients over the same time period for the TIC and NCIT-1 when NCIT-2 was in use.

Lin’s concordance correlation coefficient (CCC) was calculated to determine the agreement on temperature obtained by the devices.^21^ The value increases as a function of the nearness of the data’s reduced major axis to the line of perfect concordance (the accuracy of the data) and of the tightness of the data about its reduced major axis (the precision of the data).

The difference in the mean of each patient’s skin temperature measurement from each pair of devices was plotted against the mean of these two mean measurements to create a Bland-Altman plot,^22^ and the 95% limits of agreement (LOA) were estimated.

Analyses were conducted using mean values of repeated measurements for a participant at a specific timepoint, apart from calculations relating to modelling skin temperature change over time, reliability, and repeatability, where the original repeated measurements were used. Outlying repeated measurements were removed based on the frequency distributions of the standard deviations of repeated temperature measurements (**eFigures 1-2**).

#### Sample size

The sample size for the cohort study (N=220, allowing for 10% lost to follow-up) was determined by its primary objective to identify predictors of cellulitis recurrence.^19^ This was, therefore, the limit on this methods comparison study. Stata v18.0 software (StataCorp LLC) was used for all statistical analyses.

## Results

202 patients were included; the median age was 66 years (interquartile range, IQR 51,79), 84 (42%) were female, and 191 (95%) of white ethnicity.

For the TIC and NCIT-1, across days 1-3, missing data ranged from 16-29%, whereas day 0 data were missing for 69% (**eTable 1**). As NCIT-2 measurements were performed on fewer patients, missing data was 59-65% and 80%, respectively.

### Skin temperature over time

#### Absolute limb temperature

Across days 0 to 3 and for all devices, the mean affected limb temperature was warmer than the mean unaffected limb temperature (**Figure 1**). Including all repeated measurements in linear mixed models, the estimated day 0 affected limb temperatures were 33.06°C (95%CI 32.68 to 33.44) for the TIC, 35.22°C (34.83 to 35.61) for NCIT-1, and 36.89°C (36.56 to 37.20) for NCIT-2 (**Table 1**).

**Figure 1.**
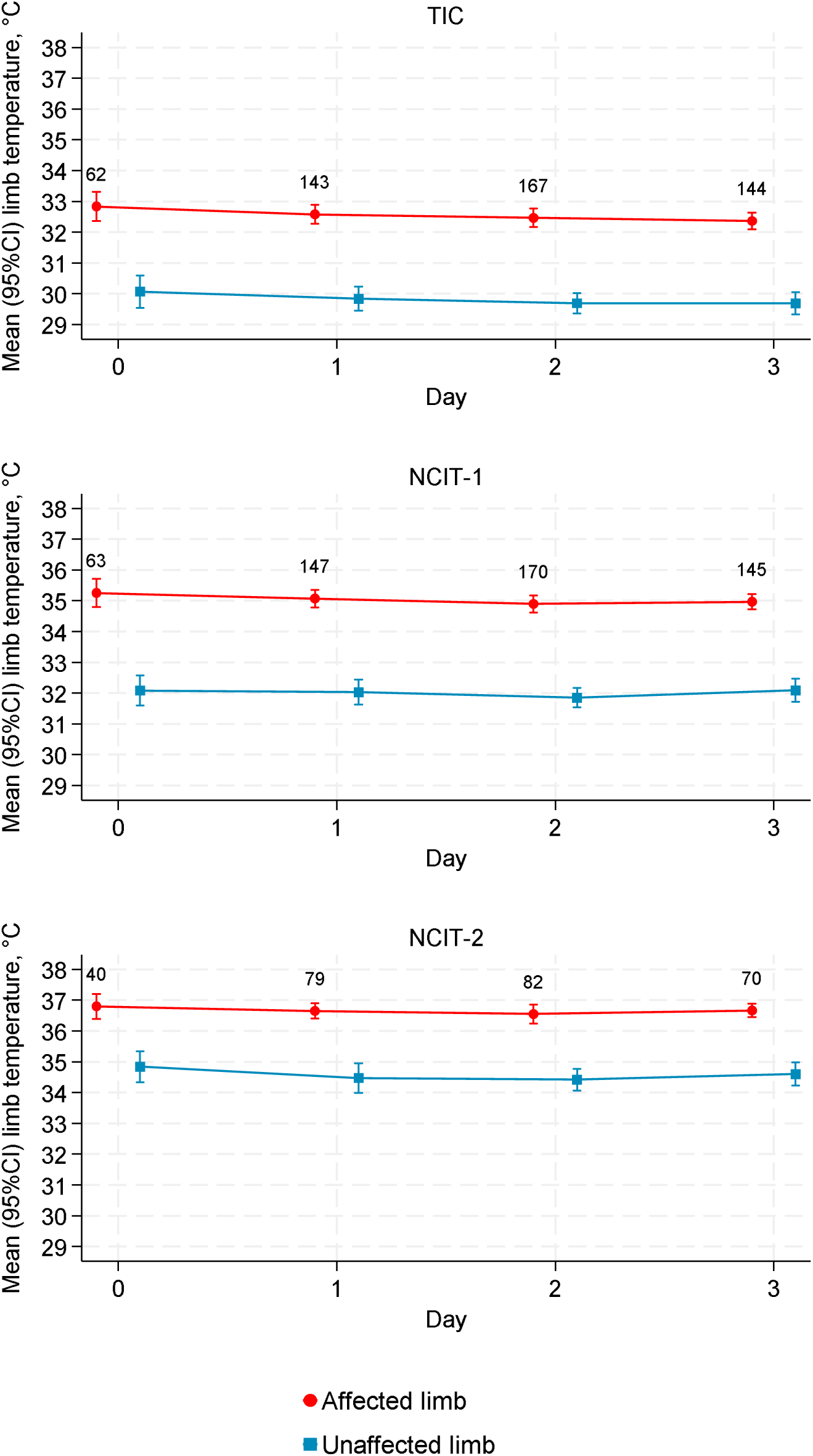
Mean (95% CIs) temperature of affected and unaffected limbs over days 0 to 3. Note: Day 0 was taken as the date of hospital antibiotic initiation. Numbers show the number of participants with data (out of a total of 202 with any data).

**Table 1.** Mean daily change in temperature in affected and unaffected limbs, and mean temperate difference between affected and unaffected limbs.

| Measurement | Device | Mean estimated temperature day 0 | 95% CI | Mean change per day (°C) | 95% CI | P value | Correlation between baseline and daily change | 95% CI | N |
| --- | --- | --- | --- | --- | --- | --- | --- | --- | --- |
| Affected limb temperature | TIC | 33.06 | 32.68 to 33.44 | -0.34 | -0.48 to -0.19 | <0.001 | -0.70 | -0.78 to -0.58 | 1,029 |
|  | NCIT-1 | 35.22 | 34.83 to 35.61 | -0.20 | -0.37 to -0.03 | 0.02 | -0.79 | -0.84 to -0.71 | 1,567 |
|  | NCIT-2 | 36.89 | 36.56 to 37.20 | -0.20 | -0.38 to -0.02 | 0.03 | -0.65 | -0.76 to -0.50 | 811 |
| Unaffected limb temperature | TIC | 29.95 | 29.48 to 30.42 | -0.11 | -0.29 to 0.07 | 0.24 | -0.79 | -0.85 to -0.71 | 1,029 |
|  | NCIT-1 | 31.85 | 31.37 to 32.32 | 0.05 | -0.15 to 0.24 | 0.63 | -0.81 | -0.86 to -0.75 | 1,563 |
|  | NCIT-2 | 34.48 | 34.00 to 34.97 | 0.02 | -0.20 to 0.23 | 0.88 | -0.74 | -0.83 to -0.62 | 807 |
| Limb temperature difference | TIC | 3.10 | 2.75 to 3.44 | -0.22 | -0.37 to -0.07 | 0.004 | -0.78 | -0.84 to -0.71 | 1,029 |
|  | NCIT-1 | 3.35 | 2.91 to 3.80 | -0.24 | -0.44 to -0.04 | 0.02 | -0.80 | -0.85 to -0.73 | 1,558 |
|  | NCIT-2 | 2.39 | 1.85 to 2.92 | -0.20 | -0.49 to 0.09 | 0.18 | -0.79 | -0.86 to -0.70 | 805 |

Note: From linear mixed models

The temperature in the affected leg decreased day by day for all devices, with the largest decrease for the TIC (−0.34°C per day, 95%CI −0.48 to −0.19, P<0.001) (**Table 1**). There was no evidence of a change in temperature of the unaffected leg per day for any device (P>0.2). In the affected leg, baseline temperature and change per day were strongly negatively correlated for all devices, i.e. limb temperatures declined the fastest in patients who started with higher limb temperatures.

#### Limb temperature difference

Across days 0 to 3, the largest mean temperature differences were recorded by NCIT-1 and the smallest by NCIT-2 (**Figure 2**). Including all repeated measurements in linear mixed models, the estimated day 0 limb temperature differences were 3.10°C (95%CI 2.75 to 3.44) for the TIC, 3.35°C (2.91 to 3.80) for NCIT-1, and 2.39°C (1.85 to 2.92) for NCIT-2 (**Table 1**).

**Figure 2.**
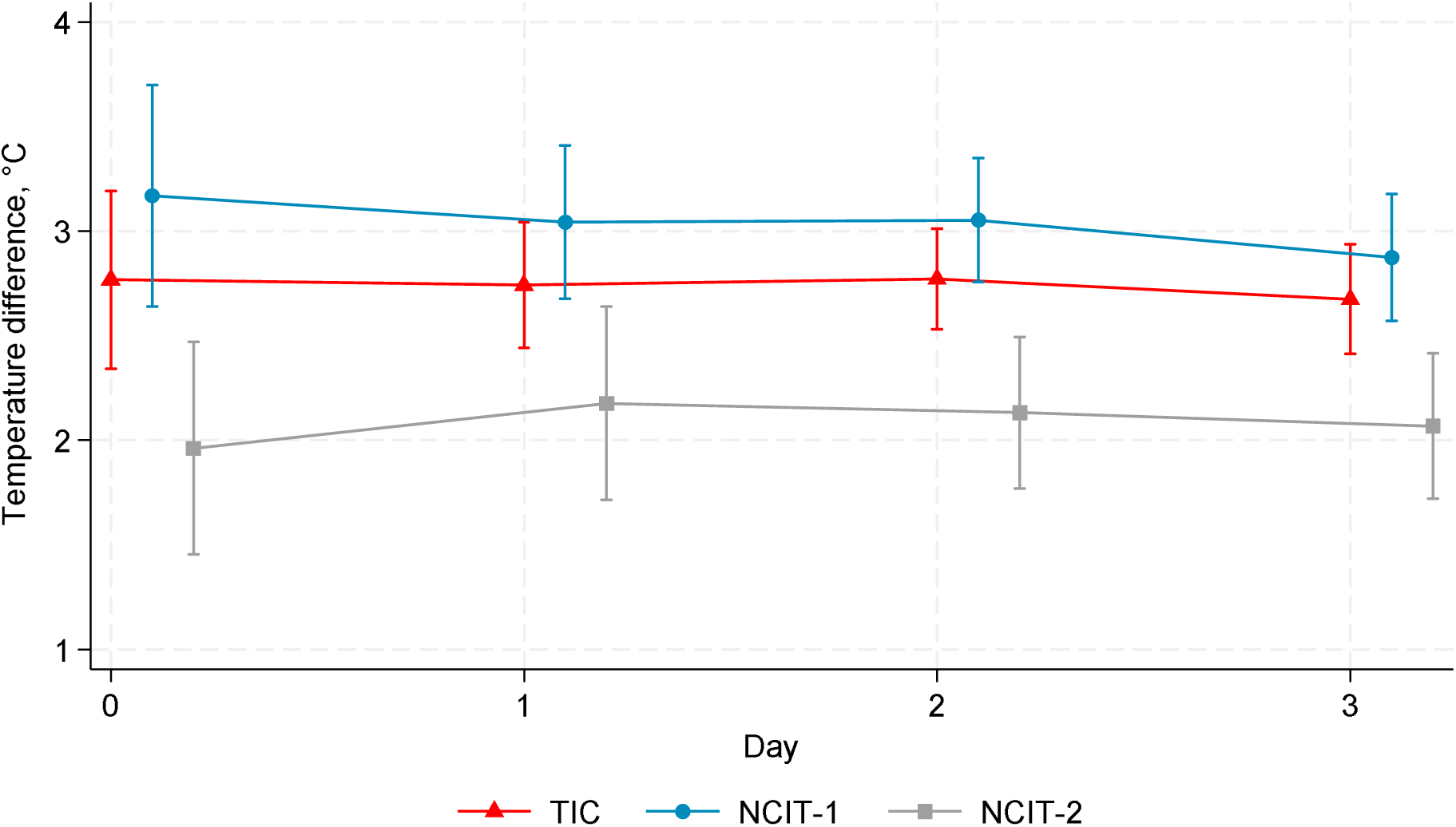
Mean (95%CIs) limb temperature difference over days 0 to 3.

The limb temperature difference decreased day by day only for the TIC and NCIT-1 (**Table 1**), by −0.22°C (95%CI −0.37 to −0.07, P=0.004) and −0.24°C (95%CI −0.44 to −0.04, P=0.02), respectively. Again, baseline temperature difference and change per day were strongly negatively correlated, i.e. limb temperature differences declined the fastest in patients who started with the greatest limb temperature differences.

### Device comparison

#### Reliability and repeatability

All three devices had excellent reliability with one-way random effects, absolute agreement, single rater intraclass correlation coefficients for repeated affected and unaffected limb temperature measurements of ≥0.98 (**eTable 2**).

Repeatability varied significantly between devices and was consistently better for affected limb measurements (**eTable 2**). Repeatability was best for NCIT-2 (0.34°C 95%CI 0.30-0.37), worse for NCIT-1 (0.54°C (0.50-0.58), and worst for the TIC (0.68°C (0.61-0.75) in the affected limb measurements. A sensitivity analysis restricted to the study period when all three devices were in use produced comparable results (**eTable 2**).

#### Agreement for affected limb temperature

The three devices recorded markedly different temperatures. The TIC recorded, on average, temperatures that were lower than NCIT-1 and NCIT-2, by −2.52°C (95% LOA −5.47 to 0.43) and −4.67°C (95% LOA −6.53 to −2.82), respectively (**Figure 3 & eFigure 3**). The largest mean differences and the lowest Lin’s concordance coefficient were observed between the TIC and NCIT-2.

**Figure 3.**
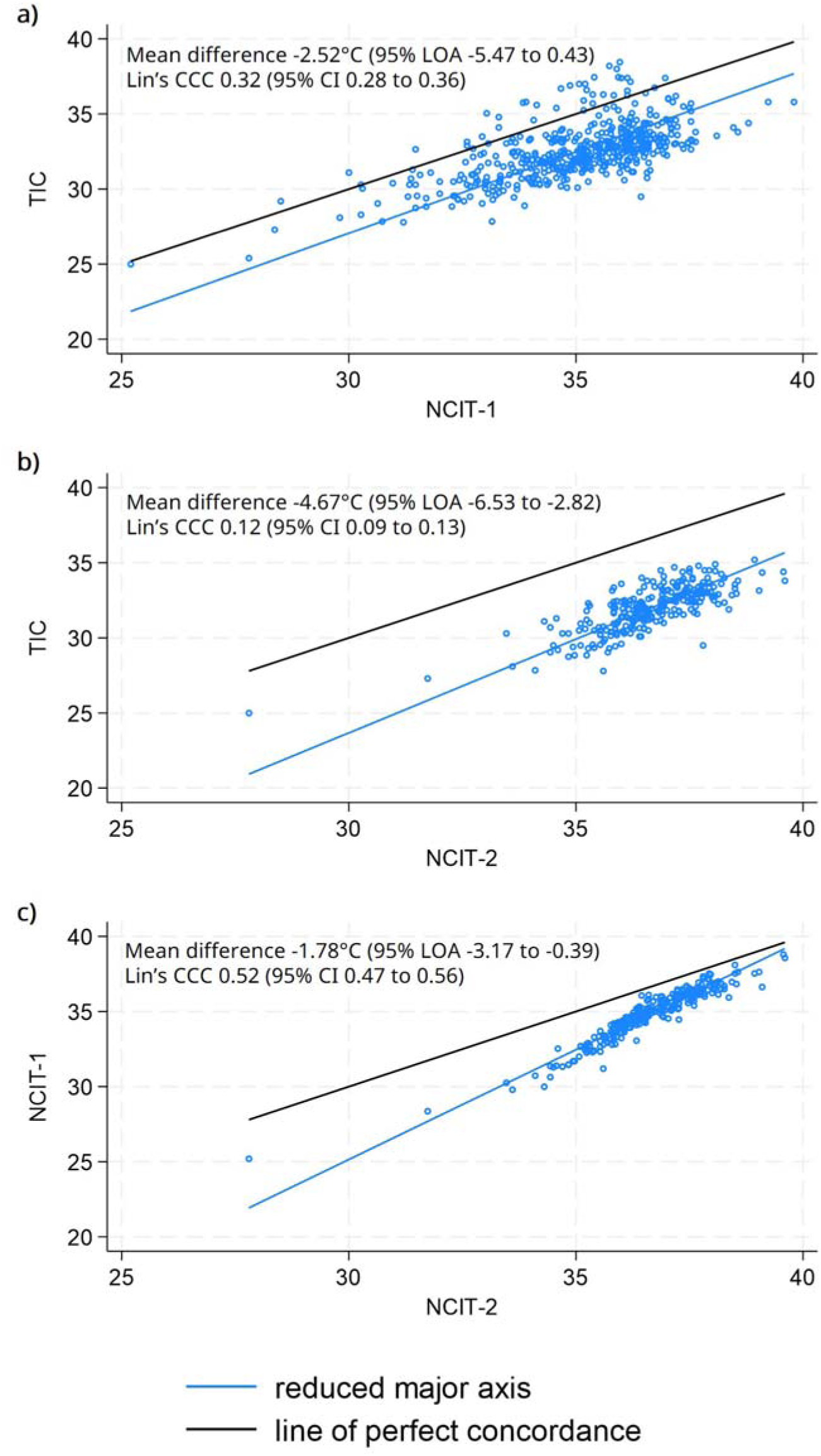
Comparison of measurements of affected limb temperature (a) TIC vs NCIT-1 (b) TIC vs NCIT-2 (c) NCIT-1 vs NCIT-2. Note: A mean difference of −2.52°C (95% LOA −5.47 to 0.43) for the TIC vs. NCIT-1 means that, on average, the TIC measures 2.52°C lower than NCIT-1 and that 95% of the measurement differences between devices will be between −5.47°C to 0.43°C.

When comparing the TIC and NCIT-2 (and NCIT-1 vs NCIT-2), the methods did not agree equally through the range of temperature measurements; as the mean temperature decreased, the difference between the measurements increased, indicating proportional bias (**Figure 3 & eFigure 3**). No such trend was observed comparing the TIC to NCIT-1, suggesting that NCIT-2 might overestimate to a greater extent at lower temperatures.

#### Agreement for limb temperature difference

There was greater agreement (higher Lin’s concordance coefficients) for limb temperature difference than affected limb temperatures (**Figure 4 & eFigure 4**). The TIC recorded, on average, lower limb temperature differences than NCIT-1 by −0.27 (95% LOA −2.65 to 2.10) and higher limb temperature differences than NCIT-2 by 0.64 (95% LOA −1.54 to 2.82). However, the mean difference was greater for the latter comparison.

**Figure 4.**
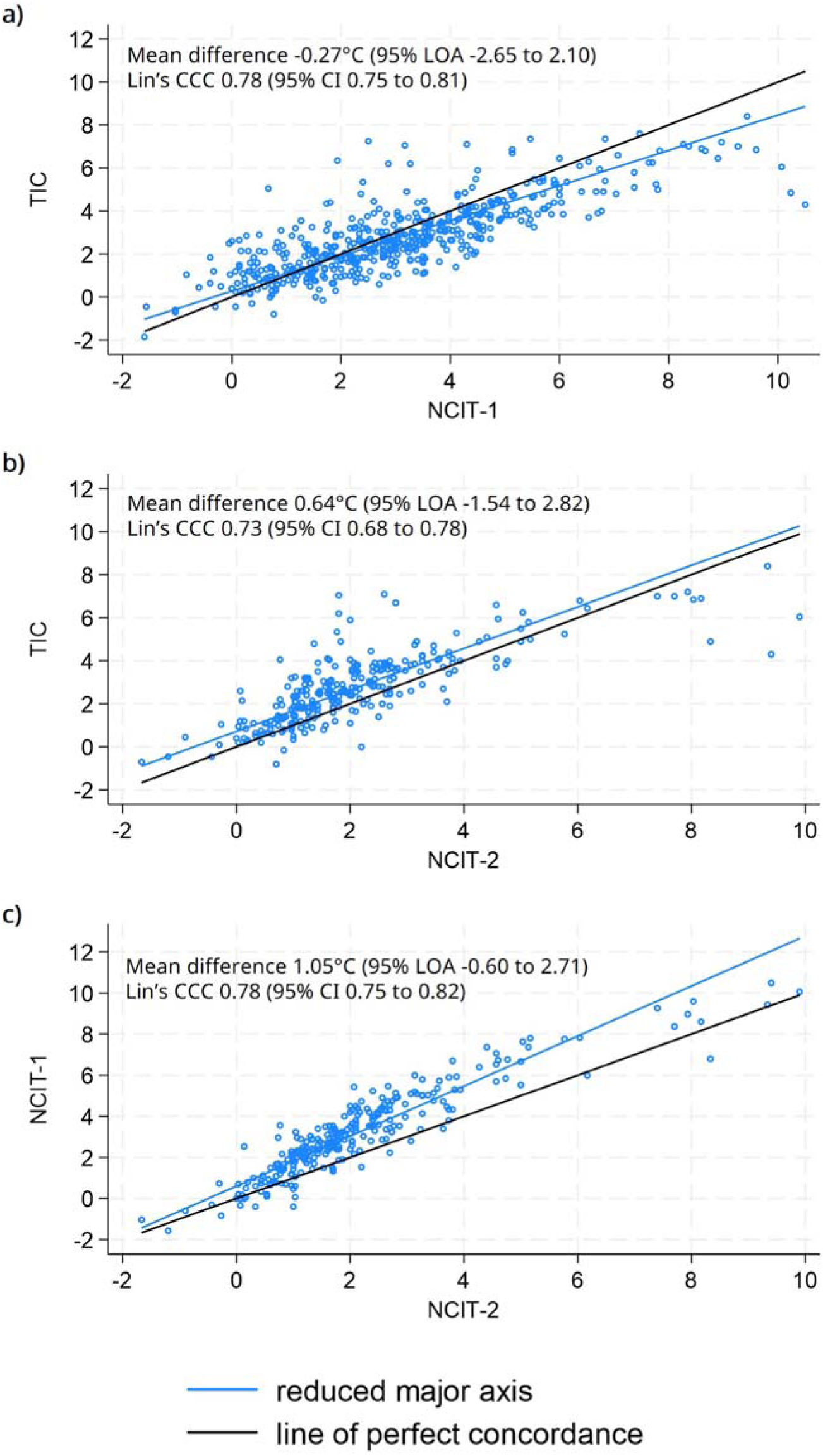
Comparison of measurements of limb temperature difference (a) TIC vs NCIT-1 (b) TIC vs NCIT-2 (c) NCIT-1 vs NCIT-2.

Lin’s concordance coefficient was higher for the TIC vs NCIT-1 comparison, 0.78 (95% CI, 0.75-0.81), than TIC vs NCIT-2, 0.73 (95% CI 0.68-0.78). As expected, the evidence of proportional bias seen for affected limb temperature between the TIC and NCIT-2 was removed when measuring limb temperature difference, given the extent and direction of bias would be the same for both limbs.

## Discussion

In patients with lower limb cellulitis prescribed antibiotics from hospital settings, the affected limb remained hotter than the unaffected limb from day 0 to day 3. Baseline affected limb temperatures varied between 33.1 to 36.9°C and limb temperature differences between 2.4 to 3.4°C, depending on the measurement device. All devices recorded a significant reduction in affected limb temperature per day, but only the TIC and NCIT-1 recorded significant reductions in limb temperature difference per day. NCIT-2 consistently recorded the smallest differences in limb temperatures, and there was evidence of proportional bias in this device, most likely due to NCIT-2 overestimating temperatures. Therefore, we would not recommend further investigating NCIT-2 to diagnose or monitor skin temperature in cellulitis.

The repeatability for the TIC was the poorest, but we only performed two repeated measurements for this device due to the time taken (~2 minutes per measurement and upload), whereas three were performed for the NCITs, which could explain the difference. Of note, we report anecdotally that the visualised temperature of the leg with the TIC appeared to fluctuate slightly in a pulse-like manner for some patients, which was assumed to be due to the patient’s actual pulse and may also explain the poorer repeatability.

Limb temperature difference diagnostic thresholds for cellulitis set in previous studies range between 0.47-0.80°C;^11,13–15^ we could not assess the actual proportion of patients meeting this threshold at baseline due to missing data. However, we did find that limb temperature difference values were relatively similar between the TIC and NCIT-1, suggesting that they could be used interchangeably. However, the most recent and largest (N=204) study suggested a threshold of 31.2°C in the affected limb temperature (not temperature difference) achieved the highest sensitivity and negative predictive value to diagnose cellulitis.^18^ Mean baseline affected limb temperatures in our study did exceed this threshold, but as measured absolute temperatures varied so markedly between devices, we would not recommend interchangeable use for absolute temperatures.

Two previous studies have used NCITs to monitor limb temperature over time.^10,12^ Montalto and colleagues followed 63 patients with cellulitis being treated by a ‘Hospital in the Home’ service and reported a mean baseline limb temperature difference of 3.5°C (95% CI 3.0-3.9), consistent with our findings from the TIC and NCIT-1.^10^ Williams *et al.* analysed trial data from 247 patients with mild to moderate lower limb cellulitis and reported a median baseline limb temperature difference of 2.3°C (IQR 1.2, 3.6).^12^ While not directly comparable to our data, this slightly lower value may be explained by patients in their study having milder disease severity or using a different NCIT.

In terms of temperature changes over time, Montalto *et al*. reported a mean reduction of 2.4°C (95% CI 1.9-3.0, P<0.001) in limb temperature between baseline and discharge (range 3 to 16 nights, not standardised across patients); we cannot easily compare our results with these varying timescales. Williams *et al.* estimated a mean reduction in affected limb temperature of 1.4°C (95% CI,1.0-1.8, P<0.001) in trial patients at the day 5 follow-up.^12^ Given we only measured through day 3, this is broadly consistent with our estimate of −0.34 °C decrease per day measured by the TIC.

Only one study has monitored cellulitis over time using thermal imaging.^17^ Amendola *et al.* analysed thermal images using a fiducial marker to estimate the relative size of the affected area. They found daily reductions in severity (i.e., normalised temperature) and scale (i.e., affected area with elevated temperature), but the unit changes reported are difficult to interpret and cannot be compared with our findings. In our study, we found the largest estimated decrease in affected temperature per day for measurements taken by the TIC, possibly suggesting it is the most sensitive measurement tool of the three evaluated. However, NCIT-1 measured a similar decrease to the TIC when estimating the change in limb temperature difference over time.

Compared with the three other studies that have measured limb temperature over time in cellulitis, our study monitored early clinical response to treatment daily in the largest population. We described limb temperature progression in patients with more severe disease than those included in the other studies (trial participants with mild to moderate cellulitis and those eligible for ‘Hospital in the Home’ treatment). We also directly compared TICs vs NCITs for measuring limb temperature in cellulitis, which no previous study has done.

Main study limitations include the fact that missing data on day 0 was high due to admissions occurring outside of the study’s working hours; our use of linear mixed models enabled estimation of temperature decreases over time, assuming data are missing at random (which includes dependence on previous/subsequent values). While attempts were made to minimise the impact of environmental factors on limb temperature, the prior positioning of the patient (lying in bed, sitting up) and the room temperature between participants and days could not be controlled. However, this reflects the constraints that would be present on a thermometry device operating in real-world clinical settings, and the unaffected limb temperature should have controlled for these factors.

Regarding generalisability, the study was only conducted at two hospital sites, although these were a mixture of tertiary care and district general services. Our findings only apply to patients with lower limb cellulitis and may not be generalisable to less severe cases treated in primary care. Finally, most patients were of white ethnicity, so our findings must be confirmed for patients with different skin tones.

From our findings and due to a lack of a gold standard, there is no clear superior method between the TIC and NCIT-1. NCITs are cheaper, require less training, and do not require a smartphone or additional software. However, our study clarifies that NCITs’ measurement capabilities differ widely and that these devices cannot be used interchangeably. Furthermore, the largest estimated decrease in affected temperature per day was measured by the TIC, and the most experience in temperature measurement in cellulitis comes from TICs^13–18^ compared to NCITs.^10–12^ Also, if more advanced analyses of thermal images (e.g. monitoring change in affected areas over time) prove more useful, these devices may hold more promise. Indeed, Amendola *et al*. found that areas of warmth may either extend or decrease before changes in oedema or skin colour either present or recede.^17^

TICs may also provide other advantages. In our study, as reported by Amendola *et al*.,^17^ patients were keen to view their daily thermal images to track the progress of their infection. In addition, many clinicians in our study wanted to view the regular photographs we had taken from the previous days, as their shift patterns meant most of them had never examined the patient they were reviewing.

Future research should explore the potential for using TICs to diagnose cellulitis in people with darker skin tones, where perceiving colour changes may be more difficult. It should also determine the clinical utility of potential earlier diagnosis and earlier indications of therapeutic success or failure, which might be offered by thermal imaging. If such findings are prognostic, further work should ensure that these techniques are accessible and interpretable in clinical settings in real-time.

## Data Availability

The data underlying this article will be shared on reasonable request to the corresponding author.

## Author contributions

All authors contributed substantially to the design and interpretation of data, drafting the manuscript and providing final approval of the version to be published. All authors agree to be accountable for all aspects of the work in ensuring that questions related to the accuracy or integrity of any part of the work are appropriately investigated and resolved. ELAC and ASW were responsible for data analysis.

## Author access to data

ELAC had full access to all the data in the study and takes responsibility for the integrity of the data and the accuracy of the data analysis.

