## Supplementary material for "Assessing skin temperature over time in a cohort of lower limb cellulitis: A methods comparison study"

### Supplementary materials

### Supplementary methods

#### Study design and population

Further exclusion criteria were patients who:

- received antibiotic therapy for another indication that was anticipated to continue for longer than the antibiotic treatment for cellulitis and that, in the judgement of the investigator, would have impacted the study assessments.
- required a surgical procedure to treat their infection (i.e. debridement of suspected necrotising skin / soft tissue infection).
- in the judgement of the investigator, did not have a clear diagnosis of cellulitis (to enable the exclusion of infections, such as severe/deep diabetic foot infection, which may be loosely labelled as cellulitis but treated with different guideline antibiotic agents and durations)
- lacked capacity to give informed consent to participate.
- were receiving end-of-life care.
- were already involved in a CTIMP of relevance to the treatment of their cellulitis.
- were unlikely, in the opinion of the investigator, to comply with study procedures.

#### Devices

- NCIT-1) The device was set to “Surface” (as opposed to “Body”) mode.
- NCIT-2) The device was set to the “ORAL” (as opposed to “RECTAL” or “AX”) and doctor “doct” mode, which the manual states allow for “scanning the skin temperature of different body areas in order to detect possible inflammations…or any other problem which may cause an alteration of the surface skin temperature”. The “Air – not” mode was selected as the device was not used in any environment with high air-conditioning.

#### Procedures

For the TIC, the point of maximal temperature on the affected lesion was determined by localising the warmest point on the thermal image. For the NCITs, determination of the point of maximal temperature involved taking five ‘test’ readings at different points of the lesion.

The TIC was held approximately 50cm away from the affected area and captured both the affected and non-affected limbs. NCIT-1 was held approximately 5 cm perpendicular to the surface of the skin. NCIT-2 is designed so that a point of light on the skin indicates how far away to hold the device, and it was held perpendicular to the skin until the single point of light appeared (not two separate or blurred points of light, indicating that it was too close or far away, respectively).

Other data collected in the cohort study included patient demographics, comorbidities, admission details, vital signs, physical examination findings, laboratory tests (including microbiological samples), and antibiotic use. The primary outcome of the cohort study was recurrence within 90 days, defined as the initiation of new antibiotic treatment in primary care or a hospital setting for cellulitis at the same site.

### Supplementary Tables

#### eTable 1: Missing temperature measurement data (N=202)

| **Day** | **TIC** | **NCIT-1** | **NCIT-2** |
| --- | --- | --- | --- |
| **0** | 140 (69%) | 139 (69%) | 162 (80%) |
| **1** | 59 (29%) | 55 (27%) | 123 (61%) |
| **2** | 35 (17%) | 32 (16%) | 120 (59%) |
| **3** | 58 (29%) | 57 (28%) | 132 (65%) |

#### eTable 2: Reliability and repeatability of the three devices

| **Device** | **Limb** | **Intraclass correlation coefficient**  **(95% CI)** | **Repeatability coefficient**  **(95% CI)** | **N** | **Sensitivity analysis:**  **Repeatability coefficient**  **(95% CI)** | **N** |
| --- | --- | --- | --- | --- | --- | --- |
| **TIC** | Affected | 0.98 (0.98-0.99) | 0.68 (0.61-0.75) | 513 | 0.65 (0.57-0.72) | 268 |
|  | Unaffected | 0.98 (0.98-0.98) | 0.82 (0.75-0.90) | 513 | 0.74 (0.65-0.83) | 268 |
| **NCIT-1** | Affected | 0.99 (0.98-0.99) | 0.54 (0.50-0.58) | 518 | 0.51 (0.46-0.56) | 265 |
|  | Unaffected | 0.99 (0.99-0.99) | 0.64 (0.60-0.68) | 514 | 0.61 (0.55-0.67) | 262 |
| **NCIT-2** | Affected | 0.99 (0.99-0.99) | 0.34 (0.30-0.37) | 269 | 0.34 (0.30-0.37) | 265 |
|  | Unaffected | 0.99 (0.99-0.99) | 0.51 (0.43-0.59) | 265 | 0.51 (0.43-0.59) | 261 |

Note: Two repeated measurements performed for the TIC. Three repeated measurements for the NCITs.

Intraclass correlation coefficients take values between 0 to 1, with higher values indicating better reliability. A repeatability coefficient of 0.68 means that two readings using the TIC on the same affected limb will be within 0.68°C for 95% of patients. Sensitivity analysis comparing repeatability coefficient over the same time period for NCIT-1 and the TIC that NCIT-2 was in use gave comparable results.

### Supplementary Figures

eFigure 1: Frequency distribution of the SDs of repeated affected limb temperatures for the TIC, NCIT-1, and NCIT-2; outliers with SDs greater than 1.54 (N=2), 0.85 (N=5), and 0.55 (N=2) were excluded from analyses, respectively

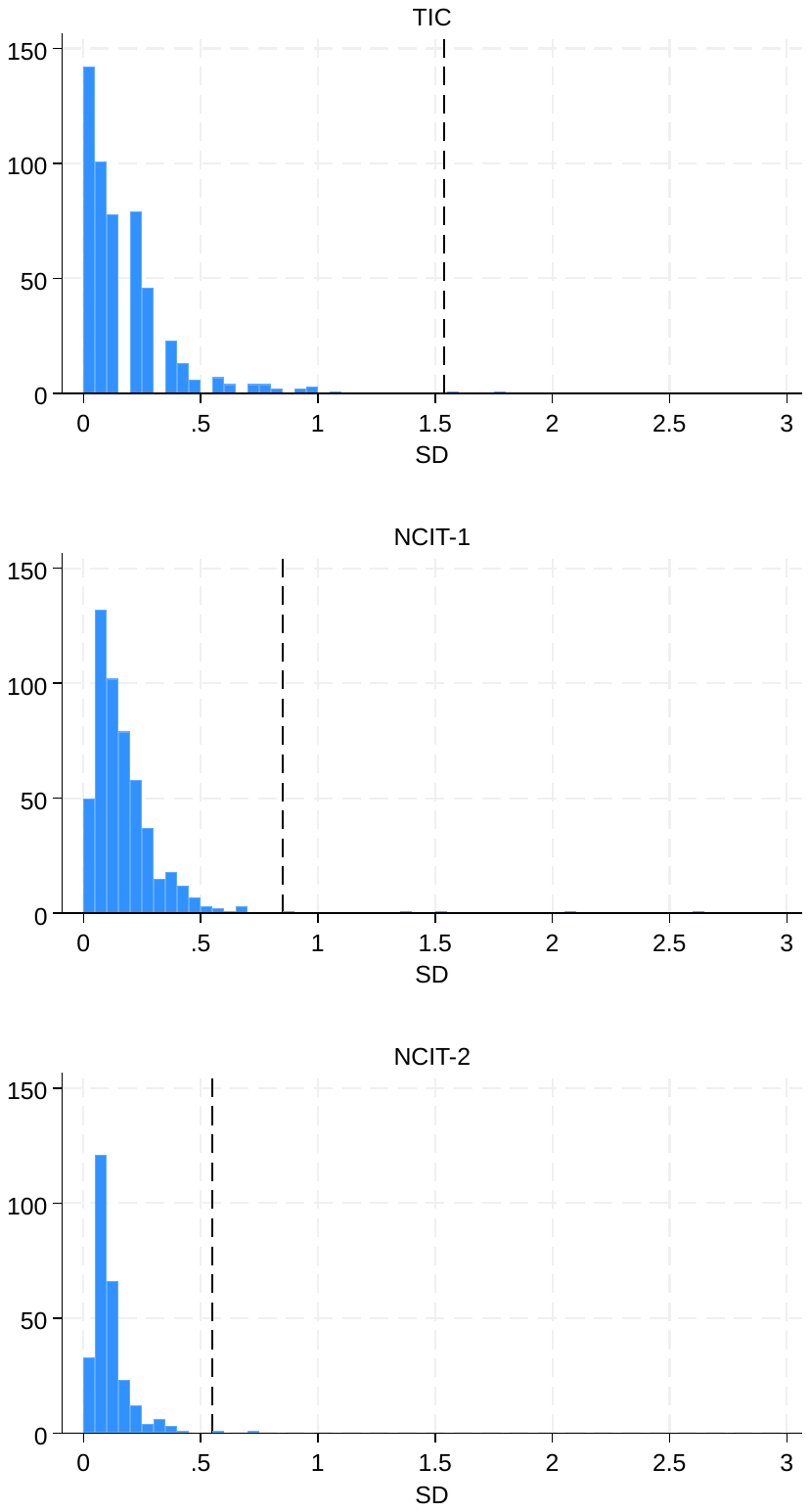

#### eFigure 2: Frequency distribution of the SDs of repeated unaffected limb temperatures for the TIC, NCIT-1, and NCIT-2; outliers with SDs greater than 1.26 (N=4), 0.89 (N=9), and 1.14 (N=6) were excluded from analysis, respectively

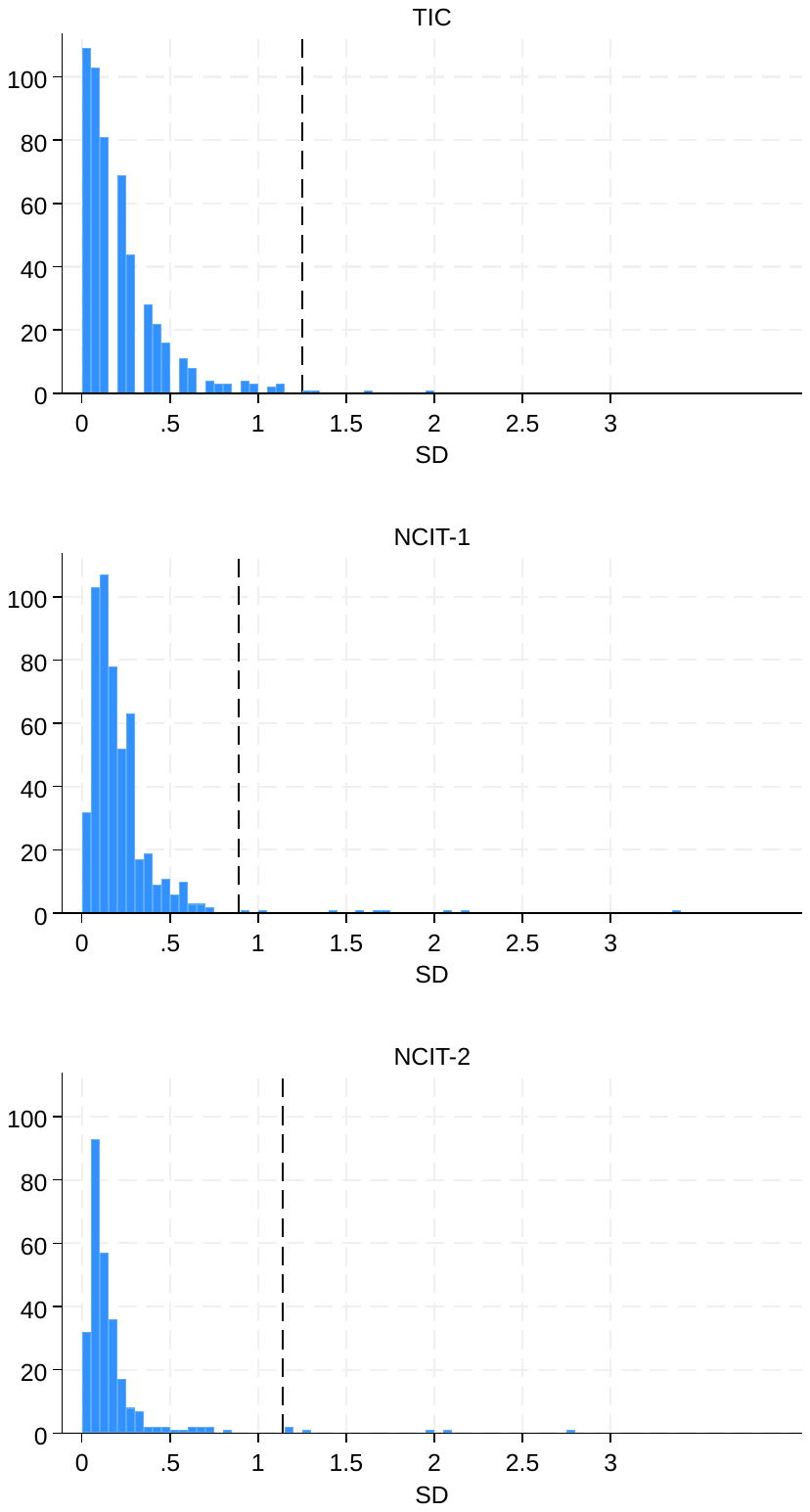

#### eFigure 3: Bland-Altman plots comparing affected limb temperatures (a) TIC vs NCIT-1 (b) TIC vs NCIT-2 (c) NCIT-1 vs NCIT-2

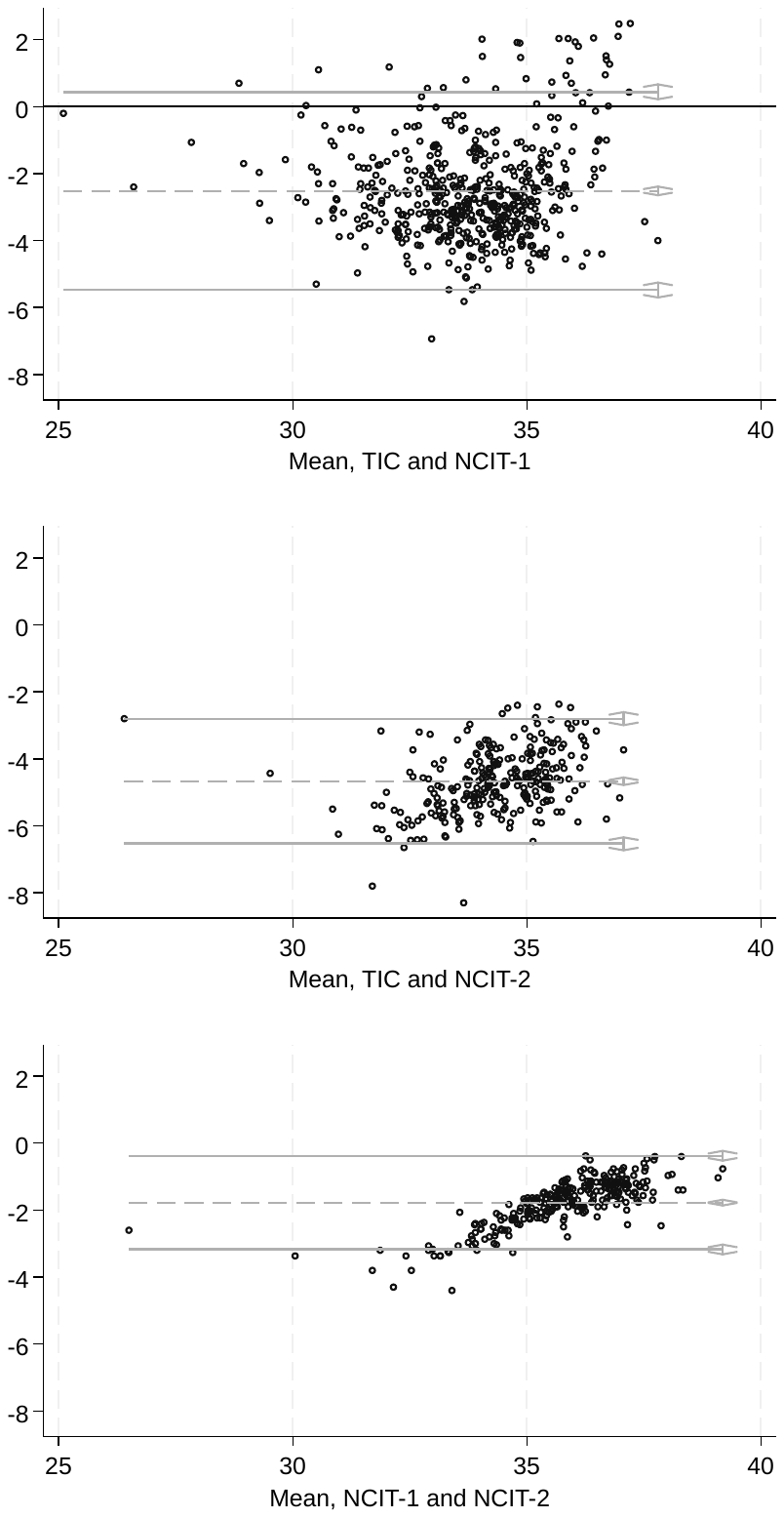

**a)**

**b)**

**c)**

Note: See eTable 3 for limits of agreement

#### eFigure 4: Bland-Altman plots comparing limb temperature differences (a) TIC vs NCIT-1 (b) TIC vs NCIT-2 (c) NCIT-1 vs NCIT-2

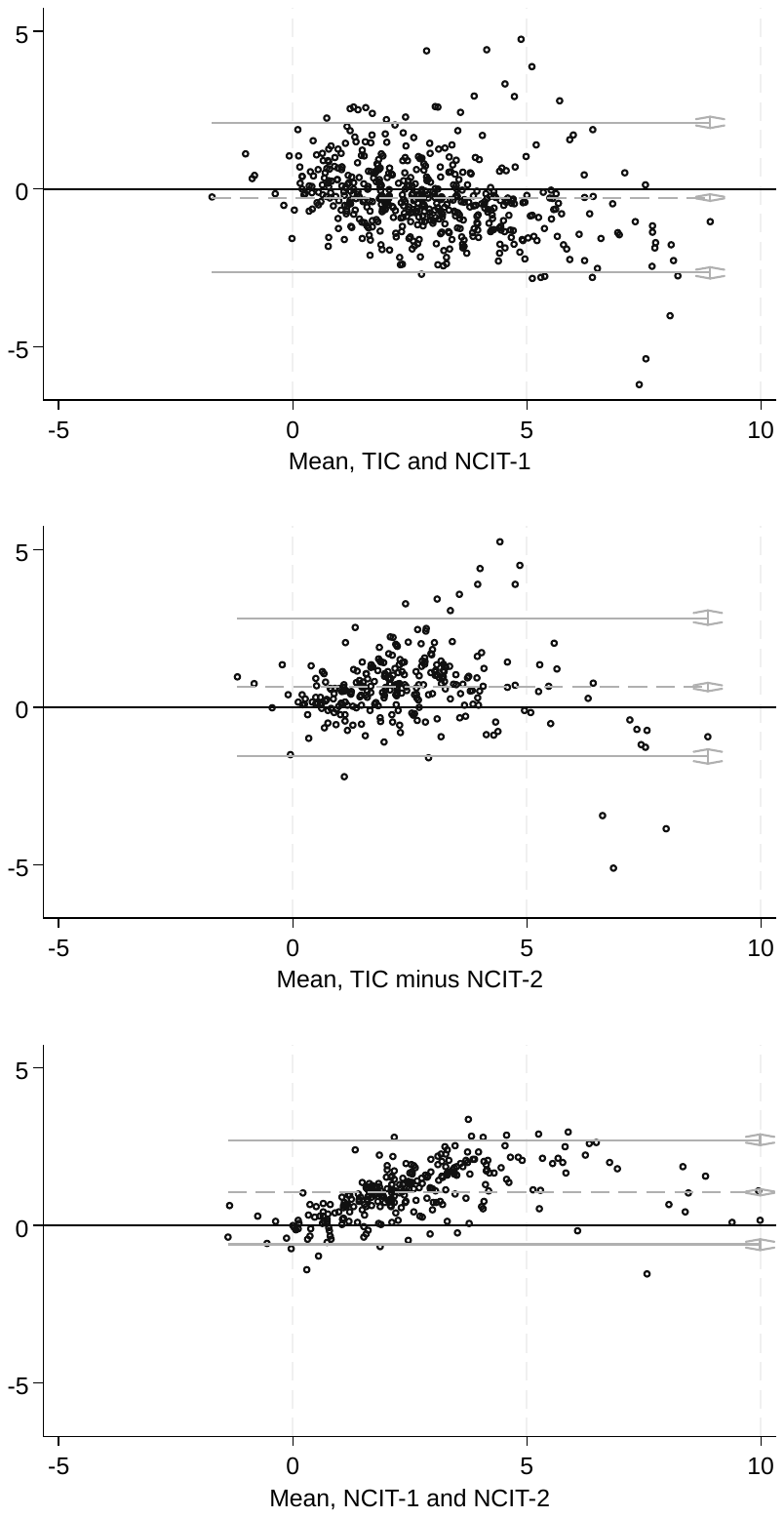

**a)**

**b)**

**c)**

Note: See eTable 3 for limits of agreement.
